# A novel use of HIV surveillance and court data to understand and improve care among a population of people with HIV experiencing criminal charges in North Carolina 2017-2020

**DOI:** 10.1101/2024.04.14.24305790

**Authors:** Elizabeth C. Arant, Andrew L. Kavee, Brad Wheeler, Bonnie E. Shook-Sa, Erika Samoff, David L. Rosen

## Abstract

**Objectives:** Objectives: To enumerate the population of people with HIV (PWH) with criminal charges and to estimate associations between charges and HIV outcomes

**Methods:** We linked statewide North Carolina criminal court records to confidential HIV records (both 2017-2020) to identify a population of defendants with diagnosed HIV. We used generalized estimating equations to examine changes in viral suppression (outcome) pre-post criminal charges (exposure), adjusting for other demographic and legal system factors.

**Results:** 9,534 PWH experienced criminal charges. Compared to others with charges, PWH were more likely to be male and report Black race. The median duration of unresolved charges was longer for PWH. When adjusting for demographic factors, the period following resolution of charges was modestly associated with an increased risk of viral suppression (aRR 1.03 (95% confidence interval 1.02-1.04) compared to the pre-charge period.

**Conclusions:** A significant portion of PWH in NC had criminal charges during a three-year period, and these charges went unresolved for a longer time than those without HIV. These preliminary findings raise questions regarding whether PWH have appropriate access to legal services.

## Introduction

Criminal system involvement in the United States (US) is common, with about 2.2 million people incarcerated in state and federal prisons and jails in a given day in 2019^1^. Formerly incarcerated people often have difficulty accessing and utilizing healthcare, resulting in negative impacts on physical and mental health^2,3^. For people with HIV (PWH) specifically, the period following prison or jail incarceration has been associated with loss of viral suppression and decreased engagement in HIV care^4,5^.

Although millions of people are incarcerated in the US annually, this population represents a fraction of all Americans who have some type of criminal record^6^. Compared to incarcerations, less is known about the effect of lower-level legal involvement. Others have hypothesized about the burden that arrests, charges, conviction, and sentencing have on physical and emotional health^7-9^. However, little is known about the impact of criminal charges in the absence of incarceration on HIV outcomes.

The goal of the current study was to enumerate the population of PWH who face charges within the criminal legal system and to estimate the association between this involvement and HIV outcomes. To do so, we used a de-identified linked database primarily composed of longitudinal North Carolina (NC) court records and confidential state HIV diagnoses and viral load records. Our central hypothesis is that being charged in the court system will be associated with declines in viral suppression, as court involvement could have a direct negative impact on disease management and serve as an indicator for behaviors that can impede disease management. This analysis represents an important step in understanding the relationship between legal system involvement and health for PWH.

## Methods

### Data source and charge definition

We utilized data from linked NC court, jail, and HIV surveillance records. This data linkage process has been described previously^10^. In summary, to identify a cohort of adult (aged ≥18) PWH in NC charged in criminal court between January 2017-February 2020, we linked the following datasets: statewide court records, jail incarceration records from 26 of 100 counties, and statewide confidential department of public health (DPH) records of all NC PWH. We obtained court records from the NC Administration Office of the Court. We derived jail incarceration histories using a technique called web-scraping, in which automated ‘bots’ collect incarceration data from public jail rosters. We then linked jail and court records using identifiers and provided these records to the NC DPH. NC DPH personnel linked these records to the state’s confidential HIV records, (using first and last name, DOB, and when available, last 4-digits of social security number), which included statewide viral load (VL) laboratory results and dates. NC DPH then provided us with linked and deidentified court and incarceration records, from which we estimated the number of adult PWH with criminal court records in all 100 NC counties. We used this de-identified database of defendants, both with and without HIV, as the data source in our analysis. The court records assign a unique identifier based on the defendant’s name and the county where a charge is initiated, therefore it is possible that one unique participant could be assigned multiple court identifiers if he/she has cases in multiple counties or is identified under a different name. For defendants with HIV, we addressed this by removing cases where a single DPH record was linked to multiple court records (n=34). For defendants without HIV, we did not have a linked DPH record and were therefore unable to eliminate redundant identifiers. Since this was a rare occurrence among PWH (i.e., < 0.5% of court records), we assumed that this was similarly rare among those without HIV and therefore unlikely to affect our findings.

In the NC court system, a defendant may face one or more criminal charges arising from a single incident. Individual charges can be adjudicated at the same time or sequentially. People may also incur new charges before existing charges are adjudicated. For the purpose of the current analysis, we considered a charge period to be a continuous period in which a person had at least one unresolved charge.

### Covariates

Both court and HIV record demographic information included DOB, race and ethnicity, and sex. HIV records included self-reported HIV transmission group and HIV VL results and dates. Of note, race and ethnicity data were reported slightly differently in the court and DPH records. Most notably, the court records included Hispanic as a race category (alongside White, Black, Other, and Unknown), whereas DPH included separate variables for race and ethnicity. Among PWH, we quantified the discordance between these two classification schemas (Supplementary Table 1). For consistency in our main analysis we reported race-ethnicity characteristics using data from the court records only.

### Outcomes

For the primary outcome of VL suppression, we adopted the NC DPH’s definition of VL suppression (<200 copies HIV RNA/ml) and un-suppression (>200 copies HIV RNA/mL or no record of a VL test within 12 months).

### Statistical Analysis

The goal of our study was to identify the population of adult diagnosed PWH in NC charged in criminal court 2017-2020 and to estimate the association between this involvement and HIV viral suppression. We used descriptive statistics to report baseline characteristics, number of discrete periods of non-overlapping charges, and duration of unresolved charge period (days). We compared these characteristics by known HIV status at the time of the first charge during the study period. For PWH who experienced a single discrete period of criminal charges during the period studied (94.1%), we calculated the proportion with viral suppression in the 12 months before initiation of and after resolution (pre-post) of criminal charges, using the VLs closest to the charge period. We included only PWH who were diagnosed at least 12 months prior to charges and had at least 12 months of follow-up following the disposition of charges. We coded PWH without a documented VL as unsuppressed. We implemented a log-binomial model using generalized estimating equations (GEE) to compare the proportion of viral suppression pre-post charges, adjusting for demographic (age, race, sex) and HIV-specific (transmission category) factors. We reported crude risk ratios (RRs), adjusted risk ratios (aRRs), and 95% confidence intervals (CI’s). We stratified the analysis of the entire population by the number of days that charges remained unresolved to determine whether our results differed by duration of unresolved charges.

Although our primary aim was to understand the relationship between having criminal charges and viral suppression in PWH in NC, we recognized that some with charges would have spent time incarcerated. Therefore, among people charged in the 26 counties for which we had jail incarceration data, we conducted a sensitivity analysis examining the association between charges and viral load suppression, comparing stratified estimates of the RR and aRR between those with and without an incarceration record.

### Ethics approval/Consent

This study was approved by the University of North Carolina at Chapel Hill institutional review board #17-0946. The IRB waived the requirements for informed consent. Data were fully anonymized before being accessed by the study team, and authors did not have access to information that could identify individual participants either during or after data collection. Data were accessed for research purposes between 5/1/2/21 and 7/1/2023.

## Results

### Full analysis

A total of 2,436,732 unique defendants experienced criminal charges in NC 2017-2020; 9,534 (0.4%) of these were PWH. Compared to those without HIV (n=2,427,198), PWH were more likely to self-identify as male (78.8% vs 69.4%). A larger number of PWH with criminal charges were Black compared to those without HIV (77.1% vs 36.8%). PWH had a median age of 41 years (interquartile range [IQR] 29-51) compared to 36 years (IQR 27-47) for those without HIV. A majority of PWH and those without HIV had a single period of criminal charges during the study period (94.1% vs 92.7%). PWH had a longer period of days with unresolved charges compared to those without HIV (median of 68 days vs 37 days). The most common mode of HIV acquisition reported at the time of diagnosis were men who have sex with men (MSM) (45.2%); 28.4% had no transmission information recorded (Table 1).

**Table 1.**
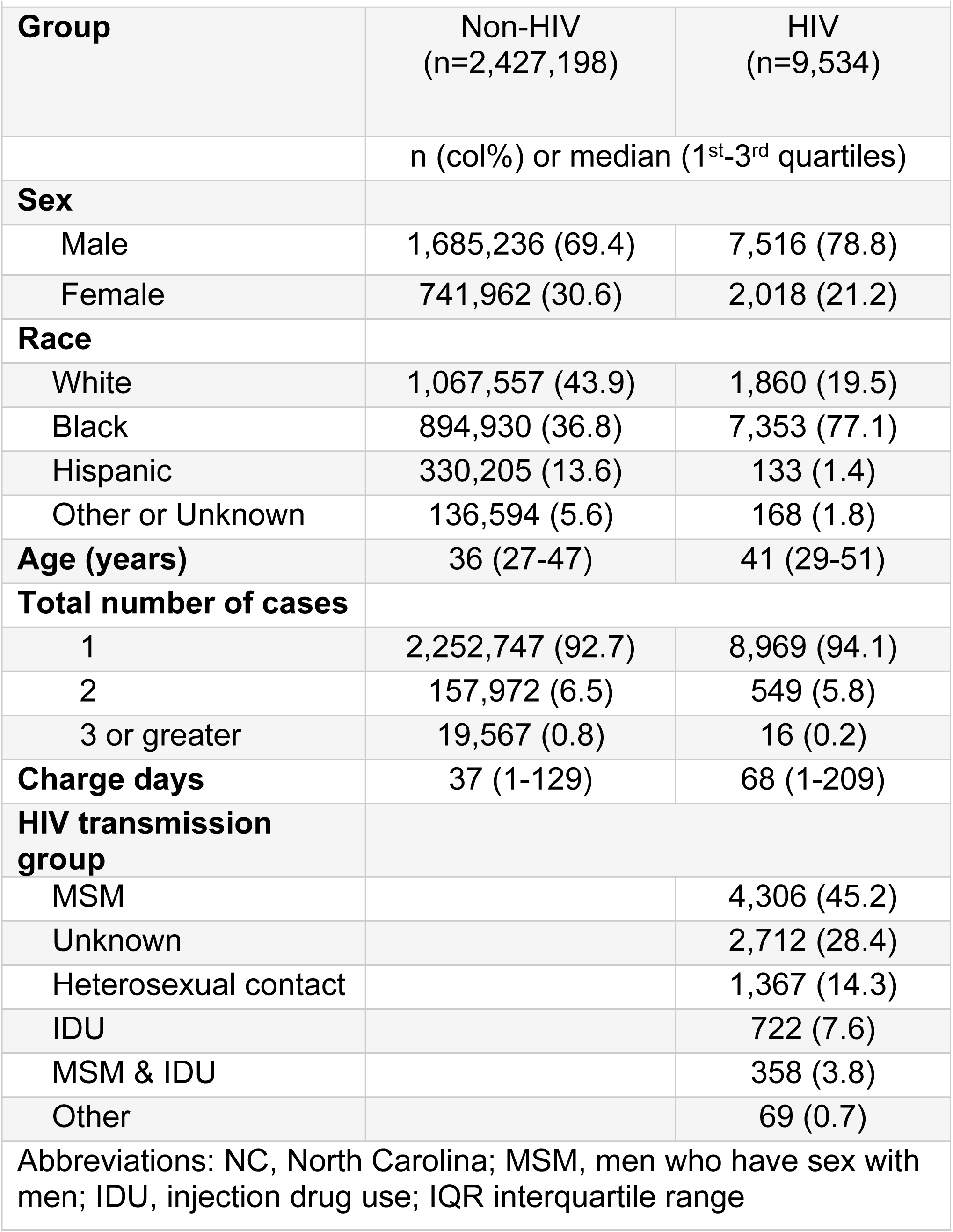
Baseline characteristics of people with and without HIV who were charged in NC criminal court, 2017-2020.

Of the 9,534 PWH with criminal charges, 3,072 (32%) were excluded from multivariable analysis, because they did not contribute 12 months of observable person-time before and after the period of criminal charges. An additional 35 (0.4%) were excluded due to missing a value for at least one covariate, leaving 6,427 (67%) available for analysis. PWH who had full follow-up periods were similar to those without based on distribution of demographic and HIV-specific characteristics in each demographic category (Supplementary Table 2). Among those with complete follow-up periods, 662 (10.2%) had a missing viral load in both the pre- or post-charge period and were therefore coded as unsuppressed.

In an unadjusted comparison, a slightly larger proportion of PWH experienced viral suppression in the post-charge period compared to the pre-charge period (72% vs 70%, *p*<0.05), corresponding to a RR of 1.03 (95% CI 1.02-1.05). When adjusting for demographic and HIV-specific factors, the period following resolution of charges was similarly associated with an increased risk of viral suppression, aRR 1.03 (95% CI, 1.02-1.04). Factors negatively associated with viral suppression in the multivariable model included Black race (aRR 0.92 [95% CI 0.90-0.95]) compared to White race and self-reported HIV acquisition via heterosexual contact (aRR 0.92 [95% CI 0.88-0.97]) or IDU (aRR 0.85 [95% CI 0.80-0.90]) compared to MSM. Factors positively associated with viral suppression included those aged 60-69 (aRR 1.27 [95% CI 1.19-1.34)] compared to those aged 18-29 (Table 2). When stratifying the analysis by duration of unresolved charges, those with the longest period of unresolved charges (300+ days, n=604 [10%]) were more likely to achieve viral suppression (RR 1.06 [95% CI, 1.01-1.12) compared to those with the shortest period of unresolved charges (0-99 days, n=4,237 [66%]), RR 1.03 [95% CI, 1.01-1.04], although the proportion with viral suppression was overall lower in both the pre- and post-charge periods among those with charges 300+ days (64.4% and 68.1%, respectively), Supplementary Table 3.

**Table 2.** Association^1^ between criminal charge period, demographics, and HIV viral suppression, NC 2017-2020 (n=6,427)^2^. Analysis including data from all NC counties.

| Characteristic | Adjusted RR (95% CI)<sup>3</sup> |
| --- | --- |
| **Time period** |  |
| 12 months pre charge |  |
| 12 months post charge | 1.03 (1.02-1.04) |
| **Age (years)** |  |
| 18-29 | Reference |
| 30-39 | 1.03 (0.98-1.08) |
| 40-49 | 1.17 (1.11-1.22) |
| 50-59 | 1.24 (1.19-1.3) |
| 60-69 | 1.27 (1.19-1.34) |
| 70 and older | 1.19 (0.99-1.43) |
| **Race/Ethnicity** |  |
| White |  |
| Black | 0.92 (0.90-0.95) |
| Hispanic | 1.05 (0.96-1.16) |
| Other | 0.95 (0.85-1.06) |
| **Sex** |  |
| Male |  |
| Female | 1.02 (0.98-1.06) |
| **HIV Transmission Category** |  |
| MSM |  |
| Heterosexual | 0.92 (0.88-0.97) |
| IDU | 0.85 (0.80-0.90) |
| MSM and IDU | 0.88 (0.82-0.95) |
| Other | 0.87 (0.69-0.91) |

<sup>1</sup> Multivariable log-binomial estimates. <sup>2</sup> Only individuals who contributed person-time to both pre- and post-charge periods and who had a single period of criminal charges were included in model. <sup>3</sup> Adjusted RRs are adjusted for all the variables in the table. Abbreviations: PWH, People Living with HIV; NC, North Carolina; RR, risk ratio; CI, confidence interval; HIV, human immunodeficiency virus; MSM, men who have sex with men; IDU, injection drug use. Definitions: Viral Suppression: <200 copies of HIV RNA per milliliter of blood

### Sensitivity analyses

To better understand whether jail incarceration affected the relationship between charge period and viral suppression, the above analyses were conducted in a subset of PWH with charges in the 26 NC counties where jail incarceration data were available. These results were stratified by jail incarceration status.

Among these counties, 157,049 people experienced criminal charges during 2017-2020, of which 1,304 (0.8%) were PWH. There were some demographic differences among this limited population of PWH and the overall population of PWH with criminal charges (Supplementary Table 4). We found that when stratifying PWH by jail incarceration status, the relationship between charge period and viral load was similar in both groups. This was true both for unadjusted analyses (RR: 0.98 [95% CI 0.89-1.08] among jail incarcerated, RR: 0.97 [95% CI 0.97 (0.99-1.09)] among those without jail incarceration) and adjusted analyses (Supplementary Table 5). We note that for all estimates, the association was not statistically significant and differed in direction from the estimated associations in our main analysis.

## Discussion

A large proportion of people in the US have some type of criminal record, and history of even a minor criminal record can have negative impacts on physical and mental health^6,8,9,11^. Most people facing criminal charges are not incarcerated or are incarcerated for brief periods. Through an innovative linkage of HIV surveillance and court data, we identified a population of diagnosed PWH with any criminal charges and examined the relationship between having criminal charges—including those with no or short incarcerations—with HIV outcomes. We found that over a three-year period nearly 10,000 PWH in NC experienced criminal charges, which in the year 2019, corresponded with about 1 in 5 PWH. The resolution of charges typically required 80% more time among PWH than among those without HIV. Contrary to our expectation, the period immediately following the resolution of charges was associated with a modest increase in the proportion with viral suppression.

This study represents one of the largest ever of legal involvement among PWH. While one study evaluated linkage to care in almost 10,000 PWH following release from US jails^12^, other studies of incarcerated or formerly incarcerated PWH have been smaller^4,5,13^. No other study has evaluated associations between criminal charges without incarceration and HIV outcomes.

Existing studies of HIV among justice-involved populations have shown that the prevalence of HIV in jails and prisons is about three to five times that of the general US population (with most HIV acquisitions occurring pre-incarceration)^14,15^.

Compared to studies of incarcerated populations, we found that the prevalence of HIV among those with criminal charges (0.4%) was similar to the prevalence of HIV in the general population^16^. Nevertheless, a relatively large number of PWH experienced charges. PWH experience a disproportional amount of social and economic inequality^17-19^, and our finding that charges among PWH took longer to resolve compared to those without HIV could be explained by PWH’s more modest resources. Other possibilities are that PWH had more serious charges that required longer to resolve, or that they were disproportionately charged in court districts that were slower to process cases. Regardless of the ultimate explanation, our findings suggest that PWH may, as a population, benefit from healthcare practice models that include a medical-legal partnership (MLP)^20^.

MLPs are healthcare delivery innovations that embed civil legal aid expertise into the health care team and focus on prevention by addressing upstream social and legal problems that affect health^21^. Such partnerships could be expanded to include criminal legal aid, particularly in locations and patient populations with frequent or prolonged legal contact. A group in Rhode Island has expanded the MLP model to include criminal cases, and they have reported positive experiences ^22^. While special considerations would need to be made for PWH given the sensitivity and stigma that an HIV diagnosis carries in society, this novel criminal MLP model serves as an example of a potential multidisciplinary support mechanism for PWH experiencing criminal charges, and such services could potentially be integrated into existing HIV services.

Racial disparities pervade the criminal justice system; Black Americans are disproportionately arrested and convicted and are given harsher sentences compared to Whites who commit the same crimes ^23,24^. There are also significant racial disparities in the HIV epidemic^18,25^. Systemic racism, HIV stigma, and economic and systemic barriers to health care that underlay disparities related to HIV care likely compound the existing racial disparities within the criminal justice system. In our study, we found a higher proportion of Black PWH with criminal charges compared to the general population of PWH in NC^16^. While not unexpected, our finding of a disproportionate number of Black defendents with HIV suggests that this compounding of racial disparities affects even early legal-system involvement for PWH. Combatting and and addressing systemic racism both in the HIV epidemic and in the criminal justice system are crucial to addressing disparities in these spaces, and HIV clinics could benefit from the support that an MLP model may provide to address upstream barriers related to systemic racism.

Our finding of a mild improvement in the proportion of viral suppression following the period of criminal charges was unexpected. While small (RR=1.03), this trend was present in both crude and adjusted analyses. Prior literature pertaining to HIV outcomes of PWH with legal involvement primarily focuses the effect of prison or jail incarcerations on HIV outcomes. Most studies show that virologic suppression improves during the prison incarceration period, likely due to access to HIV care and mental health services and a structured daily routine^26^. While HIV care and ART are less consistently available during jail incarcerations, jail lengths of stay are typically shorter (median of 3 days^27^), therefore the peri-incarceration period itself likely plays less of a role in viral suppression. Jail incarceration data were only available to us for a subset of counties. In these counties, a higher proportion of PWH with criminal charges were incarcerated compared to those without HIV (75.8% vs 67.8%). In our sensitivity analysis, the association between charge period and viral suppression was similar among those with and without record of a jail incarceration; this consistency suggests that incarceration did not modify the overall association between charge period and viral suppression. The direction of the charge period-viral suppression association in our sensitivity analysis was in the opposite direction of the association in our main analysis, suggesting that this association may vary by county.

While relatively minor offenses usually do not incur other post-sentencing sanctions, these low-level forms of justice system contact can negatively impact physical and mental health throughout the continuum of contact, particularly for those already experiencing social and health disparities^8,11^. We anticipated that the period following criminal charges would be associated with a decline in the proportion of PWH with viral suppression; instead, we found a small increase in the proportion of those with viral suppression following the period of criminal charges. It is likely that most of the criminal charges in our study were minor offenses or misdemeanors, such as traffic violations, that may have had little impact on HIV outcomes. When stratifying the analysis by duration of unresolved charges, however, those with the longest period of charges (300+ days) were more likely to achieve viral suppression compared to those with shorter charge periods. More investigation is needed to understand the relationship between type and duration of charges as they relate to HIV outcomes.

The proportion of PWH with viral suppression in our population was in between that of all PWH in NC in 2020 (66%) and those considered in care (85%)^28^. Given that a relatively small proportion (10%) of our population had missing viral load data, it may be more meaningful to compare the rate of viral suppression to the “in-care” population of PWH in NC. The justice-involved PWH in our study had a lower proportion of viral suppression compared to in-care PWH in NC, which is more consistent with our hypotheses regarding the effect of lower-level legal involvement on HIV outcomes. An alternative etiology relates to limitations of HIV surveillance data. It is possible that individuals who are still in care in another state or who have died may continue to contribute to the surveillance denominator, leading to an underestimation of the true rate of viral suppression. The NC DPH mitigates this issue using active surveillance among those originally identified as out-of-care. For example, in 2020, 26% of people originally deemed out-of-care were identified by NC DPH as living in aother state or as having died^28^. Nevertheless, unidentified out-migrations and deaths may still impact our comparisons of viral suppression among PWH with and with court involvement.

One limitation of our study is its reliance on surveillance data. Limitations of this approach have been discussed previously^29-31^. In our full analysis, about 10% of the population had missing VL data in both the pre- and post-charge periods. Prior literature suggests that, there is underreporting of VL to state surveillance systems^32^. As a result, viral suppression is underestimated. While there was considerable variation with laboratory reporting compliance in NC when the surveillance database was first created^33^, we believe that modern laboratory platforms and automated reporting have minimized missing data during the study period. Clinicians may decide to reduce VL testing frequency due to more effective and less toxic ART regimens, patient convenience, or cost^34^. Nevertheless, with our use of a full 12-months for our pre- and post-charge time frame, we believe that these factors resulted in “missing” VLs infrequently.

As discussed above, some PWH may receive VL monitoring in different states due to convenience or migration, and excluding or assuming a “worst case” HIV-related outcome for these individuals may result in an overestimation of those who are either out of care or not virally suppressed^35^. Assuming that those without VL data were not virally suppressed may underestimate the proportion of PWH with viral suppression. When excluding those with a missing VL from the analysis, our estimates were similar to estimates obtained when applying the missing data assumption, suggesting that coding those with missing VL data as unsuppressed had little impact on our results.

## Conclusions and Public Health Implications

Legal involvement, from minor offenses to arrests and charges leading to incarceration, is pervasive in the US. While more is known about the negative impact of incarceration on HIV-related outcomes for PWH, a better understanding of the impact of lower-level legal-involvement in this population is needed. In our study, a significant portion of PWH in NC had criminal charges during a three-year period, and these charges went unresolved for a longer time than those without HIV. These preliminary findings raise questions regarding whether PWH have appropriate access to legal services. Greater access to legal care could be facilitated by developing medical-legal partnerships at HIV care sites. Qualitative investigation of the experiences that PWH with criminal charges have with legal representation and their healthcare teams would strengthen our understanding about interventions that may be most beneficial to improving HIV care for this population. To better understand the relationship between the period of criminal charges and HIV care, an important next step would be to incorporate more granular details about the charge itself (type, severity) and the legal outcome of the charge in our analysis. Our investigation, while a preliminary step, reveals insightful information about an understudied population of PWH. Given that a significant number of PWH across the US likely experience criminal charges, ongoing work in this space is crucial for informing care for this population.

## Data Availability

The data are not publicly available due to their containing information that could compromise the privacy participants

---

**Supplementary Table 1.**
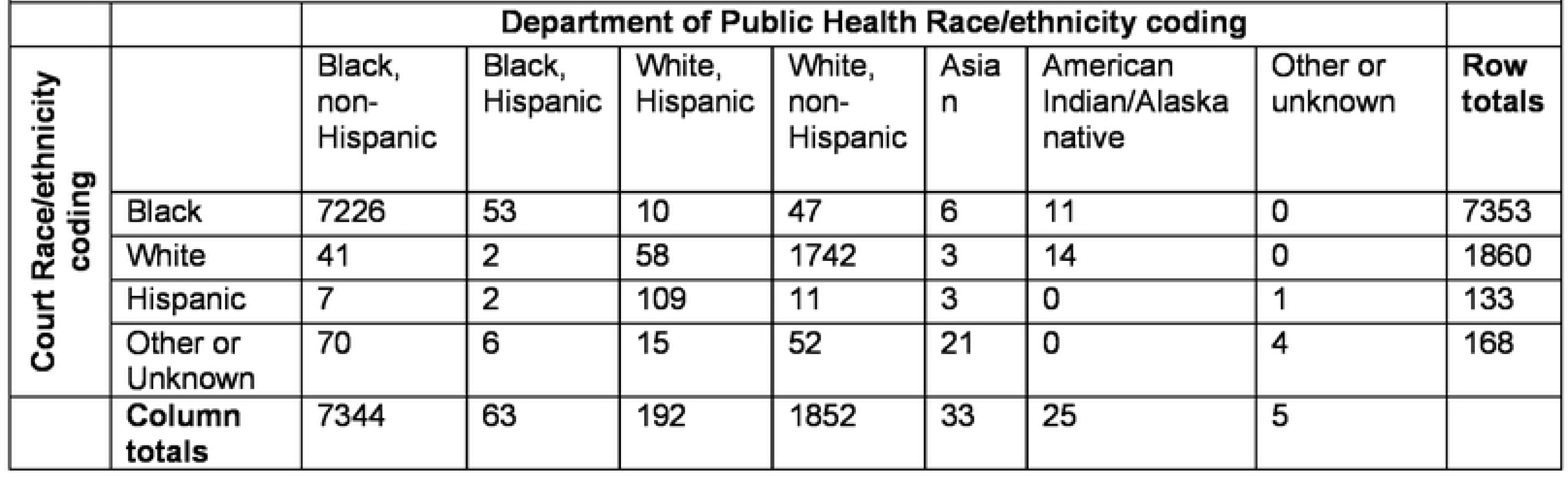
Department of public health versus Court record coding of race/ethnicity.

**Supplementary Table 2.** Baseline characteristics of people with HIV without full follow-up periods (n=2,847)

|  |  |
| --- | --- |
|  | n (col%) or median (1<sup>st</sup>-3<sup>rd</sup> quartiles) |
| **Sex** |  |
| Male | 2,228 (78.3) |
| Female | 619 (21.7) |
| **Race** |  |
| White | 568 (20.0) |
| Black | 2,204 (77.4) |
| Hispanic | 26 (0.9) |
| Other or Unknown | 49 (1.7) |
| **Age (years)** | 38 (29-50) |
| **Charge days** | 78 (1-257) |
| **HIV transmission group** |  |
| MSM | 1,305 (45.8) |
| Unknown | 771 (27.1) |
| Heterosexual contact | 410 (14.4) |
| IDU | 203 (7.1) |
| MSM & IDU | 138 (4.8) |
| Other | 20 (0.7) |
| Abbreviations: MSM, men who have sex with men; IDU, injection drug use; IQR interquartile range |  |

**Supplementary Table 3.** Association^1^ between criminal charge period and duration of unresolved charges NC 2017-2020: Results of multivariable log-binomial model (n=S,427)^2^ Analysis including data from all NC counties.

| Duration of unresolved charges in days | n (%) | % with viral suppression pre/post charge period | RR of viral suppression in post-charge period compared to pre-charge period |
| --- | --- | --- | --- |
| 0-99 | 4,237 (66) | 69.8/71.8 | 1.03 (1.01-1.04) |
| 100-199 | 984 (15) | 72.4/74.3 | 1.03 (0.90-1.06) |
| 200-299 | 592 (9) | 68.6/72.6 | 1.06 (1.01-1.10) |
| 300+ | 604 (10) | 64.4/68.1 | 1.06 (1.01-1.12) |

<sup>1</sup> Multivariable log-binomial estimates
<sup>2</sup> Only individuals who contributed person-time to both pre- and post-charge periods and who had a single period of criminal charges were included in model

**Supplementary Table 4.** Baseline characteristics of people with and without HIV who charged in criminal court in NC between 2017-2020. Data limited to 26 counties with jail incarceration data available.

|  | Jail Incarcerated and Unincarcerated |  | Jail Unincarcerated Only |  | Jail Incarcerated Only |  |
| --- | --- | --- | --- | --- | --- | --- |
| Group | Non-HIV<br>(n=155,745) | HIV<br>(n=1,304) | Non-HIV<br>(n=48,846) | HIV<br>(n=494) | Non-HIV<br>(n=105,597) | HIV<br>(n=989) |
|  | n (col%) or median (1<sup>st</sup>-3<sup>rd</sup> quartiles) |  |  |  |  |  |
| **Sex** |  |  |  |  |  |  |
| Male | 118,267 (75.9) | 1,087 (83.4) | 37,781 (77.3) | 404 (81.8) | 79,399 (75.2) | 832 (84.1) |
| Female | 37,478 (24.1) | 217 (16.6) | 11,065 (22.7) | 90 (18.2) | 26,198 (24.8) | 157 (15.9) |
| **Race** |  |  |  |  |  |  |
| White | 72,127 (46.7) | 267 (20.5) | 20,787 (42.6) | 92 (18.6) | 51,340 (48.4) | 214 (21.6) |
| Black | 72,805 (47.1) | 1,005 (77.1) | 25,556 (52.3) | 391 (79.1) | 47,997 (45.0) | 748 (75.6) |
| Hispanic | 5,582 (3.6) | 12 (0.9) | 1,415 (2.9) | 3 (0.6) | 4,177 (3.9) | 10 (1.0) |
| Other or Unknown | 3,034 (2.0) | 20 (1.5) | 1,088 (2.2) | 8 (1.6) | 2,158 (2.0) | 17 (1.7) |
| **Age (years)** | 31 (25-40) | 37 (28-49) | 31 (25-40) | 38 (28-49) | 31 (24-40) | 37 (28-48) |
| **Total number of charge periods** |  |  |  |  |  |  |
| 1 | 129,870 (84.1) | 1,301 (99.8) | 35,352 (72.4) | 492 (99.6) | 94,518 (89.5) | 986 (99.7) |
| 2 | 21,457 (13.9) | 3 (0.2) | 11,509 (23.6) | 2 (0.4) | 9,948 (9.4) | 3 (0.3) |
| 3 or greater | 3,114 (2.0) | 0 | 1,985 (4.1) | 0 | 1,129 (1.1) | 0 |
| **Charge days** | 139 (56-270) | 121 (37-277) | 91 (25-40) | 65 (2-182) | 165 (73-301) | 161 (64-309) |
| **HIV transmission group** |  |  |  |  |  |  |
| MSM |  | 506 (38.8) |  | 183 (37.0) |  | 387 (39.1) |
| Unknown |  | 414 (31.7) |  | 151 (30.6) |  | 319 (32.3) |
| Heterosexual contact |  | 175 (13.4) |  | 74 (15.0) |  | 125 (12.6) |
| IDU |  | 125 (9.6) |  | 60 (12.1) |  | 90 (9.1) |
| MSM & IDU |  | 78 (6.0) |  | 24 (4.9) |  | 63 (6.4) |
| Other |  | 6 (0.5) |  | 2 (0.4) |  | 5 (0.5) |

Abbreviations: NC, North Carolina; MSM, men who have sex with men; IDU, injection drug use; IQR interquartile range.

**Supplementary Table 5.**
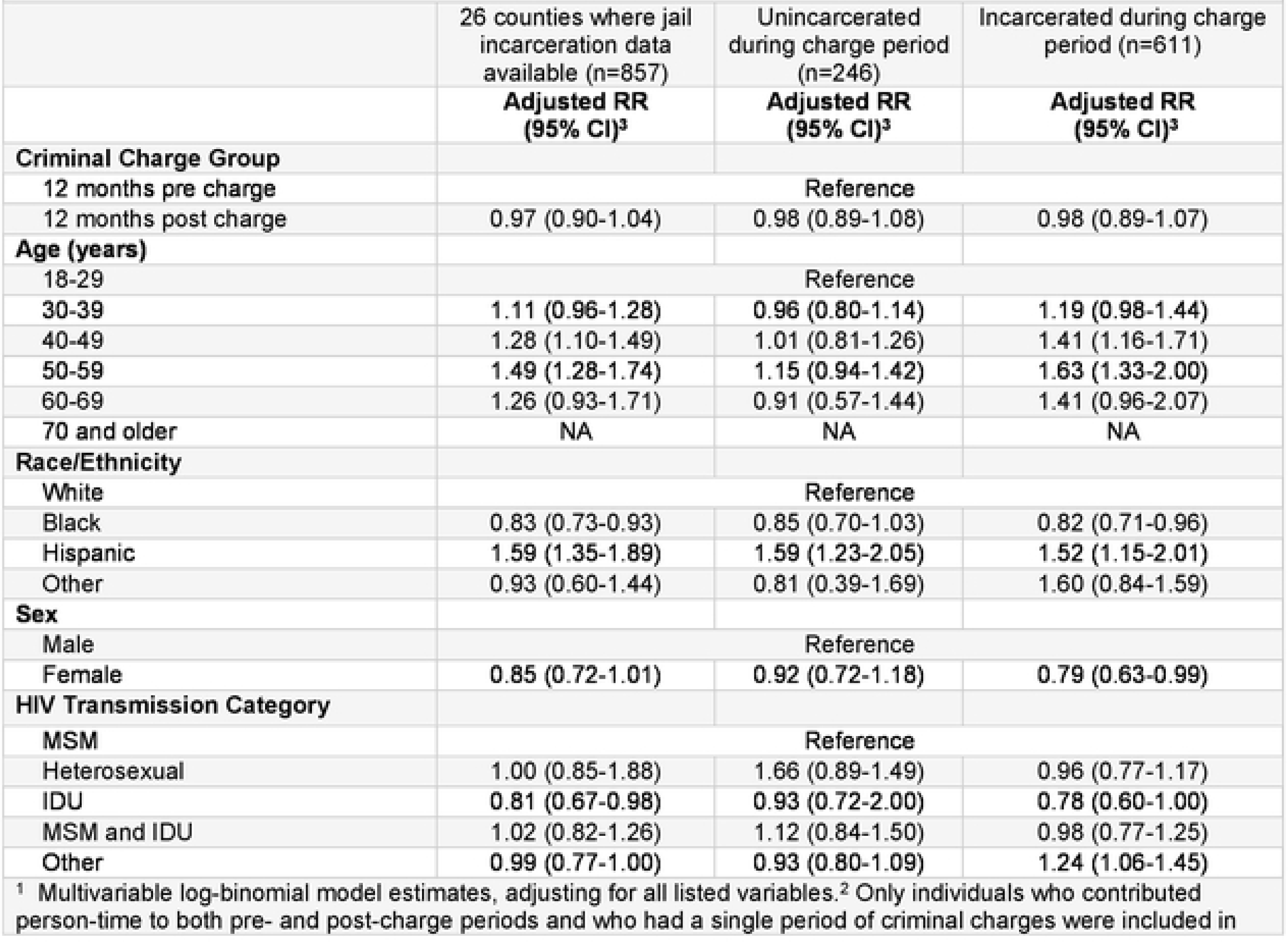
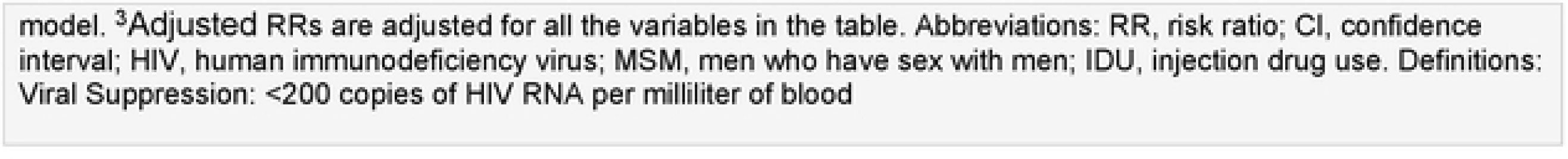
Viral suppression outcome’ among people with HIV (PWH) in NC with a single criminal charge between 2017-2020: results of multivariable log-binomial model limited to NC counties with available jail incarceration data^2^.

|  | 26 counties where jail incarceration data available (n=857) | Unincarcerated during charge period (n=246) | Incarcerated during charge period (n=611) |
| --- | --- | --- | --- |
|  | **Adjusted RR (95% CI)<sup>3</sup>** | **Adjusted RR (95% CI)<sup>3</sup>** | **Adjusted RR (95% CI)<sup>3</sup>** |
| **Criminal Charge Group** |  |  |  |
| 12 months pre charge | Reference |  |  |
| 12 months post charge | 0.97 (0.90-1.04) | 0.98 (0.89-1.08) | 0.98 (0.89-1.07) |
| **Age (years)** |  |  |  |
| 18-29 | Reference |  |  |
| 30-39 | 1.11 (0.96-1.28) | 0.96 (0.80-1.14) | 1.19 (0.98-1.44) |
| 40-49 | 1.28 (1.10-1.49) | 1.01 (0.81-1.26) | 1.41 (1.16-1.71) |
| 50-59 | 1.49 (1.28-1.74) | 1.15 (0.94-1.42) | 1.63 (1.33-2.00) |
| 60-69 | 1.26 (0.93-1.71) | 0.91 (0.57-1.44) | 1.41 (0.96-2.07) |
| 70 and older | NA | NA | NA |
| **Race/Ethnicity** |  |  |  |
| White | Reference |  |  |
| Black | 0.83 (0.73-0.93) | 0.85 (0.70-1.03) | 0.82 (0.71-0.96) |
| Hispanic | 1.59 (1.35-1.89) | 1.59 (1.23-2.05) | 1.52 (1.15-2.01) |
| Other | 0.93 (0.60-1.44) | 0.81 (0.39-1.69) | 1.60 (0.84-1.59) |
| **Sex** |  |  |  |
| Male | Reference |  |  |
| Female | 0.85 (0.72-1.01) | 0.92 (0.72-1.18) | 0.79 (0.63-0.99) |
| **HIV Transmission Category** |  |  |  |
| MSM | Reference |  |  |
| Heterosexual | 1.00 (0.85-1.88) | 1.66 (0.89-1.49) | 0.96 (0.77-1.17) |
| IDU | 0.81 (0.67-0.98) | 0.93 (0.72-2.00) | 0.78 (0.60-1.00) |
| MSM and IDU | 1.02 (0.82-1.26) | 1.12 (0.84-1.50) | 0.98 (0.77-1.25) |
| Other | 0.99 (0.77-1.00) | 0.93 (0.80-1.09) | 1.24 (1.06-1.45) |

<sup>1</sup> Multivariable log-binomial model estimates, adjusting for all listed variables.<sup>2</sup> Only individuals who contributed person-time to both pre- and post-charge periods and who had a single period of criminal charges were included in
model. <sup>3</sup>Adjusted RRs are adjusted for all the variables in the table. Abbreviations: RR, risk ratio; CI, confidence interval; HIV, human immunodeficiency virus; MSM, men who have sex with men; IDU, injection drug use. Definitions: Viral Suppression: <200 copies of HIV RNA per milliliter of blood

## References

1. Carson E. Prisoners in 2019. U.S. Department of Justice. https://bjs.ojp.gov/content/pub/pdf/p19.pdf

2. Wildeman C, Wang EA. Mass incarceration, public health, and widening inequality in the USA. Lancet. Apr 8 2017;389(10077):1464–1474. doi:10.1016/s0140-6736(17)30259-3

3. Massoglia M, Pridemore WA. Incarceration and health. Annual review of sociology. 2015;41:291.

4. Dumont DM, Brockmann B, Dickman S, Alexander N, Rich JD. Public health and the epidemic of incarceration. Annual review of public health. 2012;33:325.

5. Loeliger KB, Meyer JP, Desai MM, Ciarleglio MM, Gallagher C, Altice FL. Retention in HIV care during the 3 years following release from incarceration: A cohort study. PLoS Med. Oct 2018;15(10):e1002667. doi:10.1371/journal.pmed.1002667

6. Iroh PA, Mayo H, Nijhawan AE. The HIV Care Cascade Before, During, and After Incarceration: A Systematic Review and Data Synthesis. Am J Public Health. Jul 2015;105(7):e5–16. doi:10.2105/ajph.2015.302635

7. The Sentencing Project. Americans with Criminal Records. Accessed February 14, 2023. https://www.sentencingproject.org/app/uploads/2022/08/Americans-with-Criminal-Records-Poverty-and-Opportunity-Profile.pdf

8. Kohler-Hausmann I. Misdemeanor Justice: Control without Conviction. American Journal of Sociology. 2013;119(2):351–393. doi:10.1086/674743

9. Fernandes AD. How far up the river? Criminal justice contact and health outcomes. Social Currents. 2020;7(1):29–45.

10. Sugie NF, Turney K. Beyond Incarceration: Criminal Justice Contact and Mental Health. American Sociological Review. 2017;82(4):719–743. doi:10.1177/0003122417713188

11. Shook-Sa BE, Hudgens MG, Kavee AL, Rosen DL. Estimating the number of persons with HIV in jails via web scraping and record linkage. Journal of the Royal Statistical Society: Series A (Statistics in Society). 2022;185(S2):S270–S287. 10.1111/rssa.12909

12. Sheely A, Kneipp SM. The Effects of Collateral Consequences of Criminal Involvement on Employment, Use of Temporary Assistance for Needy Families, and Health. Women Health. 2015;55(5):548–65. doi:10.1080/03630242.2015.1022814

13. Spaulding AC, Booker CA, Freeman SH, et al. Jails, HIV Testing, and Linkage to Care Services: An Overview of the EnhanceLink Initiative. AIDS and Behavior. 2013/10/01 2013;17(2):100-107. doi:10.1007/s10461-012-0339-2

14. Baillargeon J, Giordano TP, Rich JD, et al. Accessing antiretroviral therapy following release from prison. Jama. Feb 25 2009;301(8):848–57. doi:10.1001/jama.2009.202

15. Spaulding AC, Seals RM, Page MJ, Brzozowski AK, Rhodes W, Hammett TM. HIV/AIDS among inmates of and releasees from US correctional facilities, 2006: declining share of epidemic but persistent public health opportunity. PLoS One. Nov 11 2009;4(11):e7558. doi:10.1371/journal.pone.0007558

16. HIV in Prisons, 2020--Statistical Tables. (2022).

17. North Carolina Department of Health and Human Services. 2019 North Carolina HIV Surveillance Report. Accessed January 23, 2023. https://epi.dph.ncdhhs.gov/cd/stds/figures/hiv18rpt_02042020.pdf

18. Beltran RM, Holloway IW, Hong C, et al. Social Determinants of Disease: HIV and COVID-19 Experiences. Curr HIV/AIDS Rep. Feb 2022;19(1):101–112. doi:10.1007/s11904-021-00595-6

19. Buot ML, Docena JP, Ratemo BK, et al. Beyond race and place: distal sociological determinants of HIV disparities. PLoS One. 2014;9(4):e91711. doi:10.1371/journal.pone.0091711

20. Zullo AR, Adams JW, Gantenberg JR, Marshall BDL, Howe CJ. Examining neighborhood poverty-based disparities in HIV/STI prevalence: an analysis of Add Health data. Ann Epidemiol. Nov 2019;39:8–14.e4. doi:10.1016/j.annepidem.2019.09.010

21. National Center for Medical-Legal Partnership. 2023. http://medical-legalpartnership.org/.

22. Vanjani R, Martino S, Reiger SF, et al. Physician-Public Defender Collaboration - A New Medical-Legal Partnership. N Engl J Med. Nov 19 2020;383(21):2083–2086. doi:10.1056/NEJMms2002585

23. Mauer M. Addressing Racial Disparities in Incarceration. The Prison Journal. 2011/09/01 2011;91(3_suppl):87S-101S. doi:10.1177/0032885511415227

24. NAACP. Criminal Justice Fact Sheet. Accessed March 17, 2023. https://naacp.org/resources/criminal-justice-fact-sheet

25. Centers for Disease Control and Prevention. Estimated HIV Incidence and Prevalence in the United States, 2014–2018. HIV Surveillance Report. Accessed February 14, 2023. https://www.cdc.gov/hiv/pdf/library/reports/surveillance/cdc-hiv-surveillance-supplemental-report-vol-25-1.pdf

26. Meyer JP, Cepeda J, Wu J, Trestman RL, Altice FL, Springer SA. Optimization of human immunodeficiency virus treatment during incarceration: viral suppression at the prison gate. JAMA Intern Med. May 2014;174(5):721–9. doi:10.1001/jamainternmed.2014.601

27. Camplain R, Warren M, Baldwin JA, Camplain C, Fofanov VY, Trotter RT, 2nd. Epidemiology of Incarceration: Characterizing Jail Incarceration for Public Health Research. Epidemiology. Jul 2019;30(4):561–568. doi:10.1097/ede.0000000000001021

28. Centers for Disease Control and Prevention. HIV Surveillance Supplemental Report, 2022. https://www.cdc.gov/hiv/library/reports/hiv-surveillance/vol-27-no-3/index.html

29. North Carolina Department of Health and Human Services. HIV Care Outcomes in North Carolina, 2021. Accessed April 11, 2023. https://epi.dph.ncdhhs.gov/cd/stds/figures/2021-CareOutcomesHIV.pdf

30. Powers KA, Samoff E, Weaver MA, et al. Longitudinal HIV Care Trajectories in North Carolina. J Acquir Immune Defic Syndr. Feb 1 2017;74 Suppl 2(Suppl 2):S88-s95. doi:10.1097/qai.0000000000001234

31. Rebeiro PF, Althoff KN, Lau B, et al. Laboratory Measures as Proxies for Primary Care Encounters: Implications for Quantifying Clinical Retention Among HIV-Infected Adults in North America. Am J Epidemiol. Dec 1 2015;182(11):952–60. doi:10.1093/aje/kwv181

32. Lesko CR, Sampson LA, Miller WC, et al. Measuring the HIV Care Continuum Using Public Health Surveillance Data in the United States. J Acquir Immune Defic Syndr. Dec 15 2015;70(5):489–94. doi:10.1097/qai.0000000000000788

33. Miller WC, Lesko CR, Powers KA. The HIV care cascade: simple concept, complex realization. Sex Transm Dis. Jan 2014;41(1):41–2. doi:10.1097/olq.0000000000000081

34. Gray KM, Cohen SM, Hu X, Li J, Mermin J, Hall HI. Jurisdiction level differences in HIV diagnosis, retention in care, and viral suppression in the United States. JAIDS Journal of Acquired Immune Deficiency Syndromes. 2014;65(2):129–132.

35. Department of Health and Human Services. Panel on Antiretroviral Guidelines for Adults and Adolescents. Guidelines for the Use of Antiretroviral Agents in Adults and Adolescents with HIV. Accessed January 24, 2023. https://clinicalinfo.hiv.gov/en/guidelines/adult-and-adolescent-arv

36. Buskin SE, Kent JB, Dombrowski JC, Golden MR. Migration Distorts Surveillance Estimates of Engagement in Care: Results of Public Health Investigations of Persons Who Appear to be Out of HIV Care. Sexually Transmitted Diseases. 2014;41(1):35–40. doi:10.1097/olq.0000000000000072

